# Experiences of receiving and providing maternity care during the COVID-19 Pandemic in Australia: a five-cohort cross-sectional comparison

**DOI:** 10.1101/2020.09.22.20199331

**Authors:** Zoe Bradfield, Karen Wynter, Yvonne Hauck, Vidanka Vasilevski, Lesley Kuliukas, Alyce N Wilson, Rebecca A Szabo, Caroline SE Homer, Linda Sweet

## Abstract

**Introduction:** The global COVID-19 pandemic has radically changed the way health care is delivered in many countries around the world. Evidence on the experience of those receiving or providing maternity care is important to guide practice through this challenging time.

**Methods:** A cross-sectional study was conducted in Australia. Five key stakeholder cohorts were included to explore and compare the experiences of those receiving or providing care during the COVID-19 pandemic. Women, their partners, midwives, medical practitioners and midwifery students who had received or provided maternity care from March 2020 onwards in Australia were invited to participate in an online survey. which was released between 13^th^ May and 24^th^ June 2020; a total of 3701 completed responses were received.

**Findings:** While anxiety related to COVID-19 was high among all five cohorts, there were statistically significant differences between the responses from each cohort for most survey items. Women were more likely to indicate concern about their own and family’s health and safety in relation to COVID-19 whereas midwives, doctors and midwifery students were more likely to be concerned about occupational exposure to COVID-19 through working in a health setting than those receiving care through attending these environments. Midwifery students and women’s partners were more likely to respond that they felt isolated because of the changes to the way care was provided. Despite concerns about care received or provided not meeting expectations, most respondents were satisfied with the quality of care provided, although midwives and midwifery students were less likely to agree.

**Conclusion:** This paper provides a unique exploration and comparison of experiences of receiving and providing maternity care during the COVID-19 pandemic in Australia. Findings are useful to support further service changes and future service redesign. New evidence provided offers unique insight into key stakeholders’ experiences of the rapid changes to health services.

## Introduction

The COVID-19 pandemic declared by the World Health Organization (WHO) on 11^th^ March 2020 (1) has resulted in rapid, significant and previously unprecedented changes to the way maternity services are provided around the world. These changes have impacted many stakeholders of maternity services including women, their partners and support people, midwives, medical staff and midwifery students (2).

In the global context, Australia has had lower prevalence of COVID-19 positive cases and lower mortality rates when compared to similar countries. At the time of reporting, Australia had a total of 1011 COVID-19 positive cases per million people (/mill) and 25 deaths/ mill in comparison to the United States (US) who had 17, 935 cases/mill and 550 deaths/ mill, or the United Kingdom (UK) with 4947 cases/mill and 611 deaths/mill since the pandemic started (3). Reports from countries profoundly impacted by the pandemic including Italy, China, Brazil, the UK and the US have all conveyed the need to change the way that maternity services, particularly hospital–based care for childbearing women and their families, are provided (4, 5). Australia is no exception, and there have been almost weekly updates to the policies that support clinical practice and guidelines around whom may attend which pregnancy care appointments, when, and for how long (6, 7). The speed at which health services have had to change their models of providing maternity care has impacted both those that receive maternity care (women and their partners), as well as those that provide maternity care (midwives, midwifery students and doctors).

In response to the high transmission and reproduction rates of the COVID-19 virus, the underlying objective of the rapid health service redesign in Australia has been centred on ensuring physical distancing, and reducing non-urgent contact between individuals who do not usually live in the same home (8). In addition, enhanced application of Personal Protective Equipment (PPE) including masks, face shields, gowns and gloves, as well as heightened awareness of the need for hand hygiene have been implemented in an effort to minimise the potential spread of the virus (9).

Health service models that have until recently relied on physical contact for clinical assessment and face to face provision of education, health promotion, clinical care and early parenting support have been radically transformed to be provided remotely via telephone, video calls, and with significantly shortened or in some cases, cancelled face-to-face appointments (8). Antenatal assessments have been moved to telehealth appointments using phone interviewing; with minimal contact appointments reserved for pregnancy care in later gestations, or for women who have complex health conditions (8). Face to face antenatal education programs across Australia were discontinued. This decision has impacted access to information in a format that has traditionally included women and their birth support partners, preparing them for pregnancy, labour, birth, and early parenting (10). Limitations were placed on the number of support people permitted to be present during labour and birth; in many settings, women were required to nominate only one support person which has caused concern for women and their partners (11, 12)

There is also emerging concern about the health and wellbeing of health professionals who are providing direct care. A recent study in China surveyed doctors, exploring measures of psychological stress, found that staff, particularly those in areas with higher COVID-19 prevalence, displayed high levels of psychological, emotional and physical duress (13). There are similar concerns for the welfare of students, especially midwifery students who have specific clinical experience requirements that they must fulfil in order to complete their courses (14, 15).

Whilst there has been some professional commentary (15) and early reporting of the impacts of providing maternity care during the COVID-19 pandemic from other countries (5), there is limited evidence that reports on the experience of those receiving or providing maternity care during this time in Australia, especially from multiple perspectives. There is also no published research that facilitates a comparison of the experiences and impact of the pandemic on cohorts such as women, partners and relevant health professionals during this same period.

This study aim was to explore and compare the multiple perspectives and experiences of those receiving and providing maternity care in Australia during the COVID-19 pandemic.

## Methods

A cross sectional exploratory design was used to survey five key stakeholder cohorts of maternity care in Australia. Cross sectional studies are known for their utility in collecting and measuring data at discrete points in time (16). Given the rapid changes to health service delivery in response to the COVID-19 pandemic, this methodology was considered ideal to address the identified study aim of exploring and comparing the perspectives and experiences of receiving and providing care during the pandemic. Human research ethical approval was granted by Curtin University (HRE2020-0210) with reciprocal approval issued through Deakin University (2020-175) and The University of Melbourne (2057065).

### Research Setting

There are around 315, 000 births annually in Australia(17). Maternity care is provided in the public and private settings with 25% of births nationally occurring the private sector (18). Most women receive support from a partner or designated companion (19). Midwifery care is well integrated into the health system. All women have a midwife with them when they give birth and midwifery care is provided in the hospital and community settings (18). Specialist obstetricians and physicians care for women and their babies in both public and private settings, general practitioners provide primary and some secondary maternity care (20). Midwifery students are prepared in Australian universities in a range of undergraduate and postgraduate courses with Nationally standardised minimum clinical requirements (14, 21)

### Data Collection

There was no existing validated survey tool available to facilitate the exploration and comparison of stakeholders’ experiences of maternity care during a global pandemic. The survey was developed by the research team who have content expertise in each of the cohorts represented and in survey design. The research team was intentionally formed to include midwifery researchers and academics, one obstetrician/ gynaecologist, one public health clinician and two academics with expertise in psychology including one academic with expertise in research on fathers.

The online survey was hosted on Qualtrics and was designed to compare the experiences of those receiving and those providing maternity care throughout Australia during the COVID-19 pandemic, and was guided by the WHO guidelines for respectful maternity care (19). Informed consent was indicated by participants’ selecting a check box before proceeding to complete the survey. Demographic data were collected from each cohort including which Australian state they live, work or study in, Aboriginal and Torres Strait Islander (Indigenous Australian) status, gender, language spoken at home, country of birth, and age. Eleven similarly-matched questions relating to participants’ experience of receiving or providing maternity care were asked with a 6 item Likert scale ranging from strongly agree to strongly disagree. The first four survey items asked participants about their perceptions of their personal safety as well as the safety of their families. A further six questions assessed participants’ satisfaction with care, either received or provided. Finally, participants were asked to indicate the extent to which some of their experiences turned out better than they expected.. Participants were then asked to provide 3 words that described their experiences of receiving or providing maternity care during the COVID-19 pandemic.

A full outline of the survey is available in a supplementary file. The survey was piloted with up to five stakeholders from each of the five cohorts for face validity and clarity. Minor modifications were made to layout and question sentence structure as a result of the feedback received. All pilot survey entries were then deleted before releasing the survey across Australia. The survey was distributed on 13^th^ May and closed 6 weeks later on the 24^th^ June 2020. The decision to close the survey was based on a noticeable slowing in responses.

### Recruitment

Convenience sampling was undertaken using social media. We chose social media rather than recruiting through health services as this intentionally distanced this study from any specific service or provider, potentially reducing bias in the sample and facilitating freedom of response at participants’ convenience, possibly encouraging more honest sharing of experiences. The survey was advertised on relevant Australian social media pages and in health professional newsletters with the intent of targeting each of the specific cohorts. Those who had received maternity care included women who were pregnant or had given birth since March 2020, as well as partners and other support people such as extended family, friends, doulas or others who had supported a woman in pregnancy, birth or postnatal care during the identified period. Those who had provided maternity care since the beginning of March 2020 included registered midwives, doctors involved in maternity care (which encompasses General Practitioners, Obstetricians, Neonatologists and other medical practitioners) and students of midwifery courses in Australia.

### Data Analysis

Data were imported into IBM SPSS Statistics v26. Cases for which only demographic data were provided were removed from the dataset. Demographic data and COVID-19 testing status are presented as n (%). In order to avoid low cell counts for smaller cohorts, we recoded responses to the Likert scale questions into binary variables: “Agree“ (Strongly agree, agree, somewhat agree) and “Disagree“ (Strongly disagree, disagree, somewhat disagree). Chi Squared (χ^2^) tests were performed to identify differences between cohorts on each item, comparing column proportions. Bonferroni corrections were applied to adjust for multiple tests (22).

The qualitative data (three-word responses) were analysed using NVivo 12. Word frequencies were counted; we report the top 10 most commonly occurring words for each cohort response. Word clouds are a useful way to display succinct qualitative data and have significant utility to depict and rank single word responses as participants distil their thoughts into single concepts (23). Word clouds were generated, ranking all words within the individual cohort responses. The word frequency report was generated from exact match words rather than stemmed words, to ensure that the frequency reporting directly reflected the words left by participants rather than aggregating concepts through the ‘stemmed words’ approach. For example, where ‘anxiety’ and ‘anxious’ were separately mentioned these are indicated as such. The coloured words in the centre indicate the words with the highest prevalence, moving to the bold text in the outer centre circle. Finally, the words in lighter font indicate the words that constitute the remaining descriptors in the most frequent words. This data display method provides a visual comparison of the overall stakeholder experiences which is a useful way to supplement quantitative findings (24).

## Results

Among the five cohorts, a total of 3,701 complete responses were received. There was participation from individuals in each Australian state and territory for all cohorts except for the midwifery students’ cohort which was not completed by participants from the Northern Territory or Tasmania. Surveys were completed by those from a range of ethnic backgrounds including Australian Aboriginal and Torres Strait Islander people. Participants indicated over 52 countries of birth (other than Australia) with more than 11% of the total responses from individuals who speak a language other than English at home. Of the total responses, over 13% (n= 437) of the participants had been tested for COVID-19 one or more times; only n=12 (0.32%) returned a positive test. Demographic variables across all five cohorts are presented in Fig. 1.

**Figure 1.**
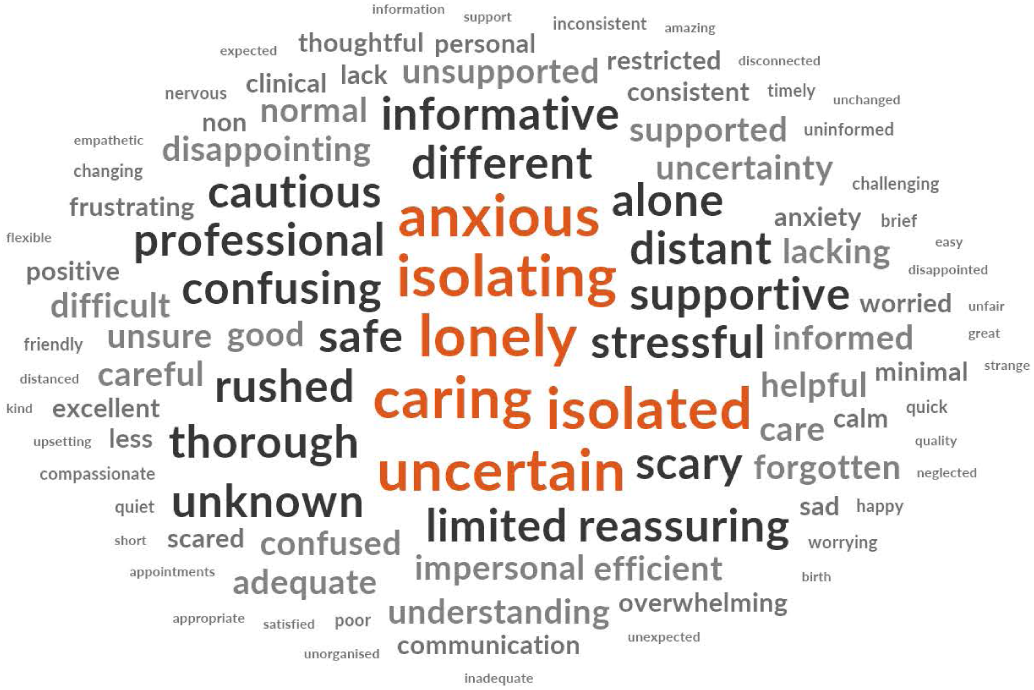
Demographic variables of cohort groups. Other^*^ Language spoken at home [*women*] includes Sinhalese, Afrikaans, Cantonese, Chinese, Dutch, German, Hindi, Italian, Malay, Mandarin, Persian, Punjabi, Sinhalese, Slovak, Tagalong, Wakhi, Nepali, Portuguese, Swedish, Russian, Urdu. Other^**^ Language spoken at home [*partners*] Dutch, Indonesian, Russian, Urdu. Other # Language spoken at home [*midwives*] Afrikaans, Arabic, Tamil. Other^β^ Language spoken at home [medical practitioners] undisclosed. Other## Language spoken at home [*midwifery students*] Vietnamese, Afrikaans, Mandarin, French. Other^◊^ Country of birth [*women*] Greece, Norway, Belgium, Bosnia & Herzegovina, Denmark, Guatemala, Indonesia, Iran, Iraq, Jordan, Kenya, Oman, Papua New Guinea, Portugal, Slovakia, Somalia, Sudan, Ukraine, Venezuela, Vietnam, Sri Lanka, Nepal, France, Russia, Sweden, Zimbabwe, Colombia, Estonia, Belarus, China, Fiji, Israel, Japan, Mexico, Netherlands, Pakistan, Poland, Samoa, Spain, Taiwan. Other^Ω^ Country of birth [*partners*] Indonesia, Belarus, Finland, India, Italy, Mexico, Netherland, NZ, Pakistan, Russia, S Africa, Zimbabwe. Other^⍰^ Country of birth [*midwives*] Ireland, Canada, Germany, Afghanistan, Austria, Czech Republic, Egypt, El Salvador, India, Kazakhstan, Malaysia, Netherlands, Philippines, Romania, Sao Tome and Principe, Singapore, Sri Lanka, Sweden, Tanzania, Turkey, US, Zambia, Zimbabwe. Other^∞^ Country of birth [medical practitioners] Malaysia, NZ, US, Canada, China, Egypt, India, Myanmar, Poland, S Africa, Tonga. Otherº Country of birth [*midwifery students*] UK, NZ, US, S Africa, China, Canada, Czech Rep, France, Singapore, Sweden.

### Abbreviations

|  |  |  |  |  |  |
| --- | --- | --- | --- | --- | --- |
| ACT | Australian Capital Territory | RNS | Rather not say | UK | United Kingdom |
| NB | Non - binary | QLD | Queensland | US | United States |
| NSW | New South Wales | SA | South Australia | VIC | Victoria |
| NT | Northern Territory | S Africa | South Africa | WA | Western Australia |
| NZ | New Zealand | Tas | Tasmania |  |  |

A comparison between the five cohorts in relation to the eleven shared survey items is presented in Fig. 2. There were statistically significant differences in the responses between the cohorts in ten of the eleven items. Women were more likely to be concerned for their own safety than each of the other cohort groups (Item 1). When considering the (potential) impact of COVID-19 on their family’s wellbeing, women were also more likely to respond that they were anxious than midwives or doctors (Item 2). Finally, when considering the health of the babies born during the pandemic, a significantly higher proportion of women were concerned for their baby’s health when compared with responses from those providing care, including midwives and doctors (Item 4).

**Figure 2.**
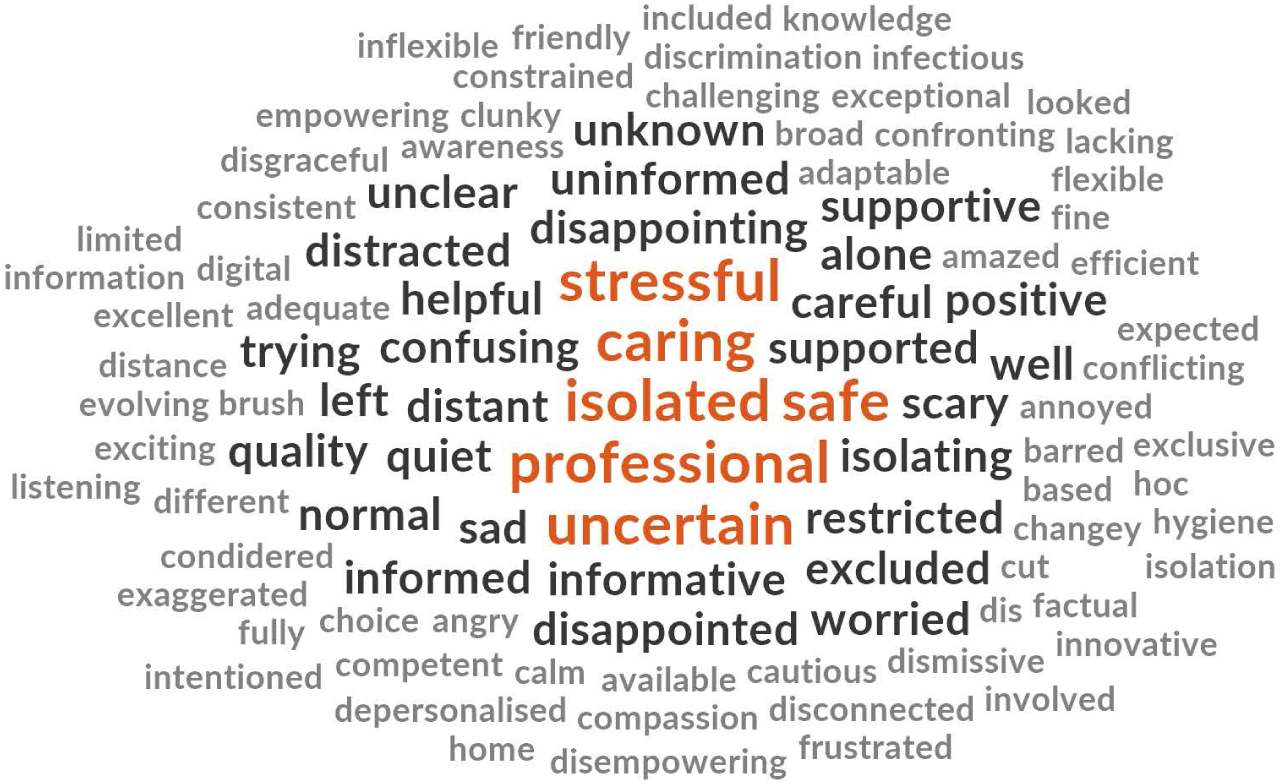
Comparison of shared items between cohort groups. Each subscript letter denotes a subset of group categories whose column proportions do not differ significantly from each other (Bonferroni adjustment applied). N/S Not significant

Regarding perceived risk of personal exposure to COVID-19 through attending or working in a hospital environment (Item 3), those providing care (midwives, doctors and midwifery students) were more likely to be concerned than women or their partners or other support people.

When asked to consider the changes in the way health services had been delivered as a result of the COVID-19 pandemic (Item 5), significantly fewer women and partners / other support people were satisfied with the changes than midwives and medical staff. Similarly, a significantly higher proportion of midwives and medical staff indicated that their professional expectations of providing maternity care during the pandemic were being met (Item 6), compared with women’s expectations of receiving care during the same period. Women’s responses to the statement in item 11 further confirmed this experience with significantly fewer women, partners / other support people and midwifery students than midwives and medical staff reporting that in comparison to their expectations, care experiences turned out better than they thought they might during the COVID-19 pandemic (Item 11). When considering the distancing measures implemented by health services during the pandemic, midwifery students were more likely to respond that they felt more isolated than women, midwives and medical professionals (Item 8).

A lower proportion of women and their partners/ other support people agreed that they were able to receive timely and clear answers to their questions about the impact of COVID-19 on their care, than midwives or doctors responding to the same question about their ability to provide timely and clear answers to women and families in their care (Item 7). Interestingly, despite this result, a higher proportion of women agreed that they were satisfied with the quality of care they received, than midwives and midwifery students who were reflecting on their satisfaction with the care they were able to provide (Item 9). This is supported with the equivocal (no significant difference) response from each cohort regarding their satisfaction with the way health services were managing risk during the pandemic (Item 10).

Participants from each cohort were asked to respond with three words that described their experiences of either receiving or providing maternity care during the COVID-19 pandemic. The words are presented in descending order of prevalence for each cohort.

**Figure 3.**
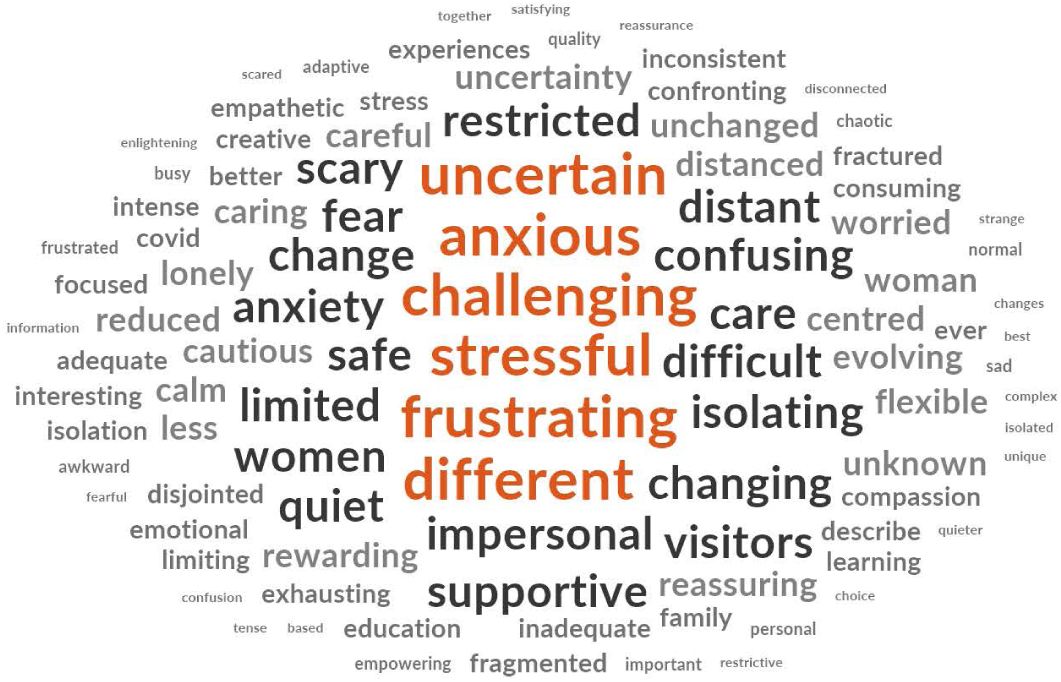
Prevalence of top ten words presented in decreasing order of frequency for each cohort.

The word clouds (Figures 4-8) are a compelling visual representation of the descriptors used by cohort participants to convey their experience of receiving or providing maternity care in Australia during the COVID-19 pandemic. Areas of similarity and divergence are apparent in this data display method which serves to support the quantitative survey findings.

**Figure 4.**
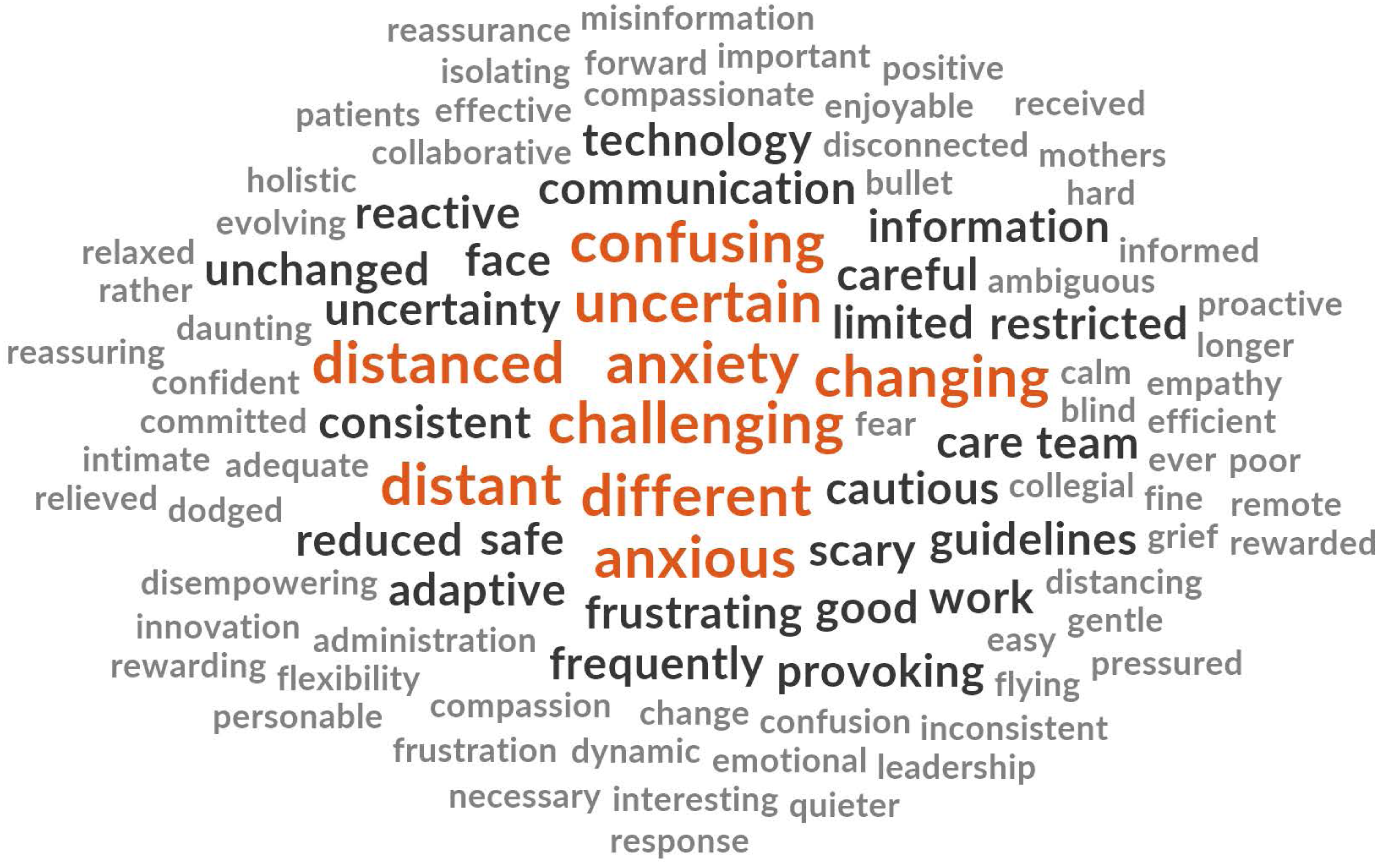
Word Cloud, Women’s Experiences of Receiving Maternity Care in Australia during the COVID-19 Pandemic.

**Figure 5.**
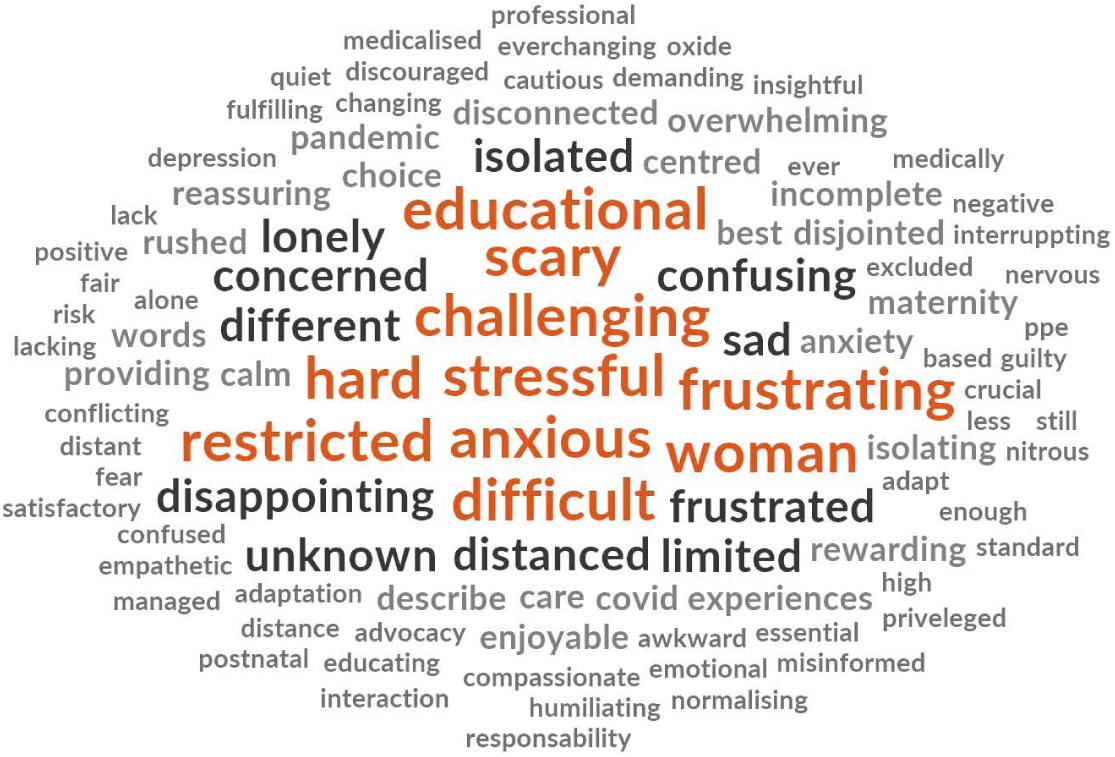
Word Cloud, Partner’s Experiences of Receiving Maternity Care in Australia during the COVID-19 Pandemic.

**Figure 6.** Word Cloud, Midwives’ Experiences of Providing Maternity Care in Australia during the COVID-19 Pandemic.

**Figure 7.** Word Cloud, Doctors’ Experiences of Providing Maternity Care in Australia during the COVID-19 Pandemic.

**Figure 8.** Word Cloud, Midwifery Students’ Experiences of Providing Maternity Care in Australia during the COVID-19 Pandemic.

## Discussion

This unique study sought to explore and compare the experiences of a range of key stakeholders in maternity care, including those who have received and provided care during the COVID-19 pandemic in Australia. Findings offer new insights into the individual and collective experience and facilitate a comparison which has highlighted areas of difference among the cohorts.

In response to the global pandemic, changes to the way maternity care is delivered have been swift, decisive and our research would suggest, disruptive. The most significant disruption was for women and their partners, who revealed that they were dissatisfied with the changes made to the way maternity care was delivered during COVID-19. A recent report from a survey of 2750 women conducted by the Australian College of Midwives (ACM) supports our findings. In the ACM report, 30% of women responded that they had considered changing their planned place of birth, 26% of women indicated that they had considered changing their care provider, 55% of these women indicated that this notion was in response to the changes made in health services such as not being allowed to have a support person attend appointments, with 36% acknowledging the reason being unable to bring their children to appointments due to physical distancing requirements (25). There was no available empirical evidence on the experience of fathers or birth support partners during COVID-19, but commentary from two Italian neonatologists note their concern about the long term impacts on the mental health and wellbeing of fathers being separated from their partners and new babies in the recent changes during the pandemic (26). The descriptions and responses from partners in our study confirm the experiences of partners’ stress, anxiety and isolation during the COVID-19 pandemic.

The findings of this research offers important insight as it reveals a divergence between consumers’ and providers’ experiences of, and satisfaction with, the way maternity health service redesign occurred during the pandemic. Whilst health professionals and midwifery students were also concerned about the changes to maternity care, it was not to the same extent indicated by women. This difference may be for a number of reasons such as health professionals’ understanding of the need to minimise risk, or due to an awareness of government regulations, and constant updates which families would not have access to. Another contributing factor may be the personal and professional divide in the nature of each cohorts’ responses. Women and their partners or support people are likely to have a deep and personal investment in their experience, and frequent changes in health care provision could understandably influence their satisfaction during periods of vulnerability. For practitioners, professional standards require flexibility and adaptability to change in the clinical environment, which may have impacted responses. Further research using in depth approaches is needed to explore the phenomenon of consumer and health practitioner responses to health service changes experienced during the COVID-19 pandemic in Australia.

When considering personal risk of exposure to COVID-19 due to presence in the hospital environment, all cohorts agreed that this risk had caused them concern. The most significant levels of worry however were reported from midwives, doctors and midwifery students. This finding may be associated with health consumers’ practice of engaging with hospitals only when necessary, whereas health professionals had no direct changes to the frequency of their attendance to hospitals as workplaces. Findings of a cross sectional survey completed by 700 maternity professionals in the UK suggested that potential lack of access to PPE, clusters of COVID-19 positive cases in local areas, and the risk of nosocomial infection may be stressors for health professionals working in maternity care (27). These assertions are supported in professional commentary offered in a reflective account of the COVID-19 experience in Kenya, Uganda and Tanzania, where midwives have expressed fear for their personal safety and potential exposure to COVID-19 (28). Further evidence is required regarding the factors contributing to practitioner concern around personal exposure and risk to families as a result of their maternity care work.

The significant changes to the provision of maternity care using telehealth and physical distancing during hospital visits had an impact on each of the study cohorts. In describing the experience, partners’ primary response was ‘isolated’. Although there is little evidence on how fathers or support partners have experienced maternity care during the pandemic, earlier research has confirmed that partners have felt isolated from inclusion in maternity care under usual circumstances (29). It is likely that the changes made to maternity care may have exacerbated this. When reflecting on feelings of isolation and loneliness experienced, midwifery students in our study were affected in significantly greater proportions than all other cohorts. Many students were prevented from attending appointments with women they had agreed to provide continuity of care throughout their pregnancy, labour and birth and into the postnatal period, a core requirement of all midwifery education throughout Australia (30). In addition, tuition changes occurred for students when universities had to move their usual face to face teaching to the online space (31). These factors are likely to have contributed to the students’ increased sense of isolation and loneliness. A cross sectional study of 972 midwifery students in Turkey revealed that they experienced high anxiety levels related to their education and clinical work as students during the pandemic (32). Although outside of the scope of this study, the authors recommend further exploration of midwifery students’ experiences with consideration to factors such as different courses, and students’ stages and progression within their course, which may have had an impact when considering implications for the wellbeing of the future midwifery workforce.

An interesting result of this study showed that midwives and doctors were more likely to feel that they could provide timely and clear answers regarding COVID-19 to women and families in their care, whereas women and partners were less likely to report they received such answers. Our findings provide a reminder of the need for clear, consistent health communication delivered in multiple modalities especially during periods of increased stress, such as those experienced in the current pandemic (33). The ACM report indicated similar findings where women described inconsistent messages and insufficient information regarding the likely impact of COVID-19 on the format of their maternity care, or impact on their babies (25).

Despite these shortcomings in communication, between 62% and 92% of each cohort agreed that their receipt or provision of maternity care during COVID-19 pandemic had turned out better than they expected; however, women, partners and midwifery students were significantly less likely to agree than midwives and doctors. Interestingly, women were more likely to indicate overall satisfaction with the care they had received than all of the other cohorts. An explanation for this result may be that although a greater proportion of women and partners or support people had indicated that their maternity care expectations were not met, overall, women were still satisfied with their care. In addition, women and partners or support people may have understood that maternity care services were doing their best to meet care needs within a rapidly evolving situation. Reports from global public health leaders indicate that in the absence of an appropriate vaccine for the COVID-19 virus, the changes made by health services will stay to a greater or lesser extent in accordance with local spread of the virus (34). In light of the findings of this study, it is timely to consider what the consequences would be if a substantial proportion of midwives and midwifery students are persistently dissatisfied with the care they can provide. A recent review suggested that staff burnout and attrition from the health care workforce are potential consequences (35). These data were was collected from May to June 2020 in the Australian pandemic experience; there is need for more research containing psychometric testing and stress inventory scales to determine the long term effect of the levels of duress that health professionals have endured, and the impact on their personal and professional identities and health.

The approach adopted in this study of considering the interconnected experience of those both receiving and providing care during the same period, provides essential knowledge urgently needed by health care leaders, policy makers, clinicians and educators. The evidence facilitates consideration for the way maternity care is continued in Australia during, and after the pandemic. Equally important, consideration of stakeholder perspectives will inform what changes should and could be made if and when the pandemic eases. It is timely and important to note the difference between the responses of consumers and providers of care so that future health service redesign may be undertaken with a holistic view by understanding what is important to consumers and providers of maternity care; and what the experiences of the current radical redesign has been.

### Strengths and Limitations

The strengths of this study lie in the exploration of a range of stakeholders’ perspectives of receiving or providing maternity care during the COVD-19 pandemic. The large sample size and timely data collection during the early peak of the pandemic has resulted in a data set that has significant utility and addresses a known gap in evidence regarding the Australian responses to maternity care during the COVID-19 pandemic, with implications that may also be relevant for consideration by other nations which have had higher prevalence rates of the COVID-19 virus.

The limitations of this research are related to the convenience sampling technique used, which involves a non-random selection of participants. The rationale for this selection has been previously described and was an important consideration for reducing the burden to a population with recognised distress during the pandemic. As is commonly found in survey-based data collection in Anglophone nations, our study was completed by participants who most often speak English at home; as such caution is advised when considering the transferability of the findings to non-English speaking groups.

## Conclusion

This study provides the first-known evidence to address the identified gap regarding multiple stakeholders’ experiences of receiving or providing maternity care during the COVID-19 pandemic in Australia. As national and international health leaders begin to consider what will be the ‘new normal’ of the way maternity care is provided, the findings highlighted in this study will contribute to an understanding of the broader human and social implications of health service redesign beyond epidemiological and financial factors.

## Data Availability

All data pertaining to the study reported in this manuscript is present within the manuscript.

## Acknowledgements

The authors would like to acknowledge the women, partners, midwives, doctors and midwifery students who participated in this study.

---

| Variables | Women<br>N=2840<br>n (%) |  | Partners<br>N=92<br>n (%) |  | Midwives<br>N=560<br>n (%) |  | Medical Practitioners<br>N=78<br>n (%) |  | Midwifery Students<br>N=131<br>n (%) |  |
| --- | --- | --- | --- | --- | --- | --- | --- | --- | --- | --- |
| Australian state<br>(live, work, study) |  |  |  |  |  |  |  |  |  |  |
| NSW | 632 (22.4%) |  | 18 (19.6%) |  | 100 (18.5%) |  | 17 (22.1%) |  | 21 (17.2%) |  |
| VIC | 842 (29.8%) |  | 27 (29.3%) |  | 185 (34.3%) |  | 24 (31.1%) |  | 38 (31.1%) |  |
| QLD | 472 (16.7%) |  | 5 (5.4%) |  | 70 (13.0%) |  | 14 (18.2%) |  | 25 (20.5%) |  |
| WA | 473 (16.7%) |  | 24 (26.1%) |  | 105 (19.5%) |  | 8 (10.4%) |  | 17 (14.0%) |  |
| SA | 194 (6.9%) |  | 7 (7.6%) |  | 47 (8.7%) |  | 5 (6.5%) |  | 20 (16.4%) |  |
| ACT | 119 (4.2%) |  | 9 (9.8%) |  | 13 (2.4%) |  | 5 (6.5%) |  | 1 (0.8%) |  |
| NT | 6 (0.2%) |  | 1 (1.1%) |  | 10 (1.8%) |  | 1 (1.3%) |  | 0 |  |
| Tasmania | 89 (3.1%) |  | 1 (1.1%) |  | 10 (1.8%) |  | 3 (3.9%) |  | 0 |  |
| Gender |  |  | Male 66 (71.7%)<br>Female 26 (28.3%) |  | Female 550 (98.4%)<br>Male 5 (0.9%)<br>RNS/NB 4 (0.7%) |  | Female 66 (84.6%)<br>Male 12 (15.4%) |  | Female 131 (100%) |  |
| Language spoken at home | English<br>Other * | 2802 (98.9%)<br>31 (1.1%) | English<br>Other ** | 88 (95.7%)<br>4 (4.3%) | English<br>Other # | 556 (99.5%)<br>3 (0.5%) | English<br>Other<sup>β</sup> | 77 (98.7%)<br>1 (1.3%) | English<br>Other<sup>##</sup> | 126 (96.2%)<br>5 (3.8%) |
| Country of birth | Australia<br>UK<br>NZ<br>S Africa<br>Ireland<br>Malaysia<br>Canada<br>India<br>Philippines<br>US<br>Germany<br>Singapore<br>Brazil<br>Italy<br>Other<sup>δ</sup> | 2445 (86.5%)<br>100 (3.5%)<br>83 (2.9%)<br>26 (0.9%)<br>15 (0.5%)<br>13 (0.5%)<br>13 (0.5%)<br>12 (0.4%)<br>11 (0.4%)<br>11 (0.4%)<br>9 (0.3%)<br>8 (0.3%)<br>5 (0.2%)<br>5 (0.2%)<br>71 (2.5%) | Australia<br>UK<br>Other<sup>Ω</sup> | 74 (80.5%)<br>5 (5.4%)<br>13 (14.1%) | Australia<br>UK<br>NZ<br>S Africa<br>Other<sup>Ⓜ</sup> | 445 (79.9%)<br>54 (9.7%)<br>18 (3.2%)<br>8 (1.4%)<br>32 (5.8%) | Australia<br>UK<br>Other<sup>∞</sup> | 58 (74.4%)<br>5 (6.4%)<br>15 (19.2%) | Australia<br>Other<sup>º</sup> | 115 (87.8%)<br>16 (12.2%) |
| Age |  |  |  |  |  |  |  |  |  |  |
| 18-25 years | 432 (15.2%) |  | 7 (7.6%) |  | 79 (14.1%) |  | 1 (1.3%) |  | 58 (44.6%) |  |
| 26-30 years | 967 (34.1%) |  | 20 (21.7%) |  | 74 (13.2%) |  | 13 (16.7%) |  | 26 (20.0%) |  |
| 31-35 years | 1035 (36.5%) |  | 29 (31.6%) |  | 78 (14.0%) |  | 13 (16.7%) |  | 22 (16.9%) |  |
| 36-40 years | 342 (12.1%) |  | 20 (21.7%) |  | 72 (12.9%) |  | 19 (24.4%) |  | 10 (7.7%) |  |
| 41-45 years | 57 (2.0%) |  | 5 (5.4%) |  | 61 (10.9%) |  | 9 (11.5%) |  | 6 (4.6%) |  |
| 46-50 years | 3 (0.1%) |  | 11 (12.0%) |  | 55 (9.8%) |  | 9 (11.5%) |  | 8 (6.2%) |  |
| 51-55 years |  |  |  |  | 63 (11.3%) |  | 5 (6.4%) |  |  |  |
| 56-60 years |  |  |  |  | 45 (8.1%) |  | 6 (7.7%) |  |  |  |
| >61 years |  |  |  |  | 32 (5.7%) |  | 3 (3.8%) |  |  |  |
| Tested for Covid19 |  |  |  |  |  |  |  |  |  |  |
| Never | 2581 (90.9%) |  | 82 (89.1%) |  | 386 (69.3%) |  | 48 (61.5%) |  | 107 (81.7%) |  |
| Once | 237 (8.3%) |  | 8 (8.7%) |  | 147 (26.4%) |  | 19 (24.4%) |  | 23 (17.5%) |  |
| Twice | 19 (0.7%) |  | 1 (1.1%) |  | 23 (4.1%) |  | 10 (12.8%) |  | 1 (0.8%) |  |
| Three or more | 2 (0.1%) |  | 1 (1.1%) |  | 1 (0.2%) |  | 1 (1.3%) |  |  |  |
| Covid19 test positive |  |  |  |  |  |  |  |  |  |  |
| No | 251 (96.5%) those tested |  | 9 (90.0%) those tested |  | 168 (98.2%) those tested |  | 30 (100%) those tested |  | 24 (100%) those tested |  |
| Yes | 1 (0.4%) |  |  |  |  |  |  |  |  |  |
| Missing | 8 (3.1%) |  | 1 (10.0%) |  | 3 (1.8%) |  |  |  |  |  |

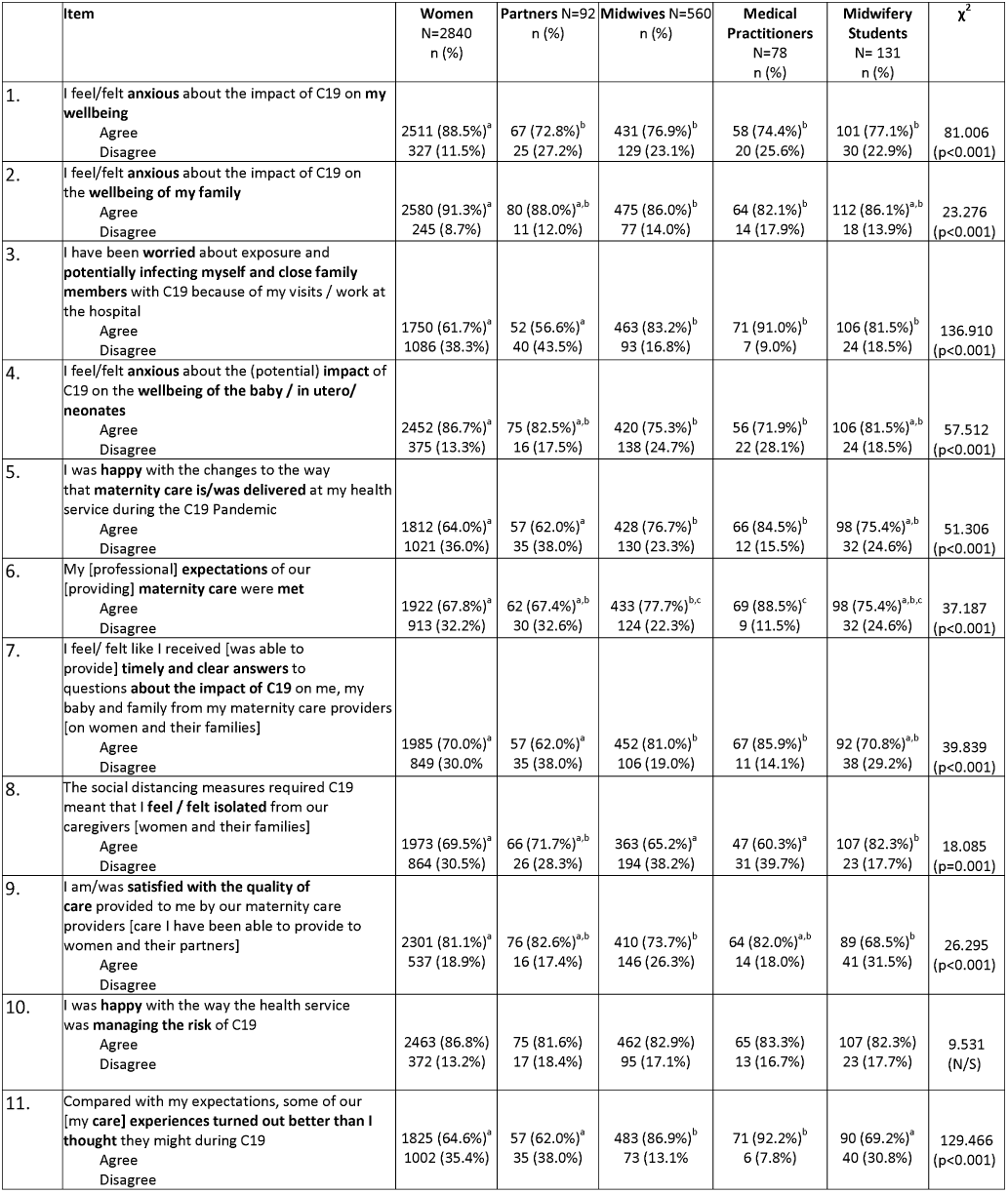

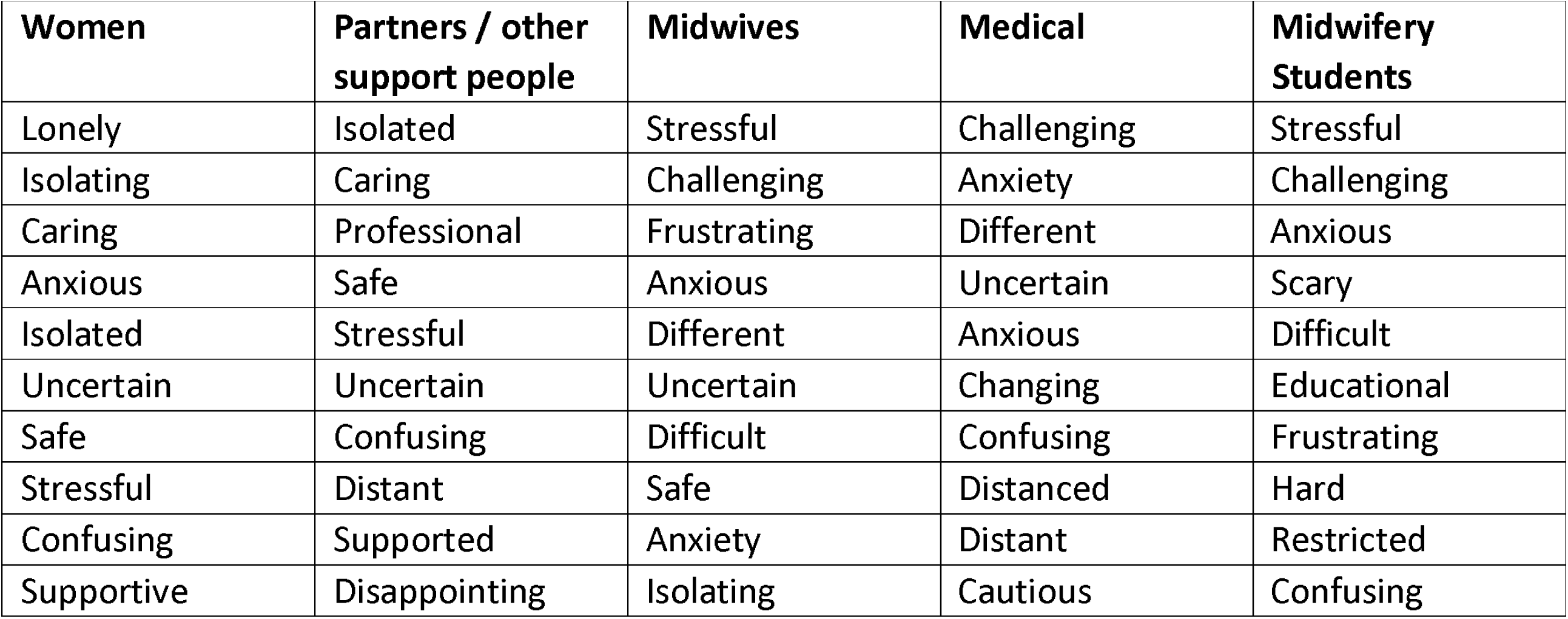

